# Three-year outcomes with fractional flow reserve- or angiography-guided multivessel percutaneous coronary intervention for myocardial infarction

**DOI:** 10.1101/2023.08.30.23294648

**Authors:** Etienne Puymirat, Guillaume Cayla, Tabassome Simon, Philippe Gabriel Steg, Gilles Montalescot, Isabelle Durand-Zaleski, Fabiola Ngaleu Siaha, Romain Gallet, Khalife Khalife, Jean-François Morelle, Pascal Motreff, Gilles Lemesle, Jean-Guillaume Dillinger, Thibault Lhermusier, Johanne Silvain, Vincent Roule, Jean-Noel Labèque, Grégoire Rangé, Grégory Ducrocq, Yves Cottin, Didier Blanchard, Anaïs Charles Nelson, Juliette Djadi-Prat, Gilles Chatellier, Nicolas Danchin, the FLOWER-MI study investigators

## Abstract

**Background:** In multivessel disease (MVD) patients with successful primary percutaneous coronary intervention (PCI) for ST-elevation myocardial infarction (STEMI), the Flow Evaluation to Guide Revascularization in Multivessel ST-Elevation Myocardial Infarction (FLOWER-MI) trial showed that a fractional flow reserve (FFR)-guided strategy was not superior to an angiography-guided strategy for treatment of non-infarct-related artery lesions regarding the one-year risk of death from any cause, MI, or unplanned hospitalization leading to urgent revascularization. The extension phase of the trial was planned using the same primary outcome to determine whether a difference in outcomes would be observed with a longer follow-up.

**Methods:** In this multicenter trial, we randomly assigned patients with STEMI and MVD with successful PCI of the infarct-related artery to receive complete revascularization guided by either FFR (n=586) or angiography (n=577).

**Results:** After 3 years, a primary outcome event occurred in 52 of 498 patients in the FFR-guided group and in 44 of 502 patients in the angiography-guided group (hazard ratio[HR], 1.19; 95% confidence interval [CI], 0.79-1.77; *P=0.4*). Death occurred in 22 patients in the FFR-guided group and in 23 in the angiography-guided group (HR, 0.96; 95% CI 0.53-1.71); nonfatal MI in 23 and 14), respectively (HR, 1.63; 95% CI 0.84-3.16); and unplanned hospitalization leading to urgent revascularization in 21 and 18 (HR, 1.15; 95% CI 0.61-2.16), respectively.

**Conclusions:** Although event rates in the trial were lower than expected, in patients with STEMI undergoing complete revascularization, an FFR-guided strategy had not a significant benefit over an angiography-guided strategy with respect to the risk of death, MI, or urgent revascularization up to 3 years. (Funded by the French Ministry of Health and Abbott; FLOWER-MI ClinicalTrials.gov number, NCT02943954.)

**CLINICAL PERSPECTIVE:** **What Is New?**

- In STEMI patients with MVD, an FFR-guided strategy is not superior to an angiography-guided strategy for treatment of non-infarct-related artery lesions regarding the risk of death from any cause, MI, or unplanned hospitalization leading to urgent revascularization at 3 years.

**What Are the Clinical Implications?**

- In patients with STEMI undergoing complete revascularization, an FFR-guided strategy had no significant benefit over an angiography-guided strategy with respect to the risk of death, MI, or urgent revascularization up to 3 years.

## INTRODUCTION

Over the last decade, several randomized trials have demonstrated that achieving complete revascularization in patients with ST-elevation myocardial infarction (STEMI) and multivessel disease (MVD) improves clinical outcomes.^1–4^ The value of a fractional flow reserve (FFR)-guided strategy for non-culprit lesions in STEMI patients is controversial. Two major studies have shown that FFR-guided complete revascularization of non-infarct related coronary lesions during the same (or a staged) percutaneous coronary intervention (PCI) procedure reduced repeat revascularizations compared with culprit lesion only revascularization.^3,4^ Based on the assumption that complete revascularization is superior to culprit-lesion-only revascularization, the Flow Evaluation to Guide Revascularization in Multivessel ST-Elevation Myocardial Infarction (FLOWER MI) randomized trial was the first study aiming at determining whether FFR-guided complete revascularization translated into better clinical outcomes, compared with angiography-guided complete revascularization in STEMI patients with MVD. The results have shown that an FFR-guided strategy was not superior to an angiography-guided strategy for reducing the risk of the composite of death from any cause, non-fatal MI, and unplanned hospitalization leading to urgent revascularization at one year.^5^ These results apparently contradict those of the subsequently published Fractional Flow Reserve vs. Angiography-Guided Strategy for Management of Non-Infarction Related Artery Stenosis in Patients with Acute Myocardial Infarction (FRAME-AMI) trial in which an FFR-guided PCI reduced a composite of death, MI, or repeat revascularization at a median follow-up of 3.5 years, compared with an angiography-guided PCI in patients with either ST-elevation or non-ST-elevation myocardial infarction (NSTEMI).^6^ The extension phase of the FLOWER MI trial was designed to determine whether a difference in outcomes would be observed beyond the initial one-year follow-up.^7^

## METHODS

### Trial design

The study design and results at one year of the FLOWER MI trial have been published previously.^5,7^ In brief, the FLOWER-MI trial was an investigator-initiated, randomized, open-label, multicenter trial with blinded end-point evaluation in which FFR-guided complete revascularization was compared to angiography-guided complete revascularization in patients with STEMI at the early stage.

All STEMI patients (≥18 years old) with successful culprit lesion PCI (primary, rescue or pharmaco-invasive; defined as TIMI flow ≥2 and residual stenosis <30%) were eligible for enrollment if the non-infarct-related coronary arteries (IRA) (i.e., major epicardial coronary artery or their major side branches ≥2.0 mm in diameter) showed ≥1 lesion with a stenosis ≥50% in diameter by visual assessment, judged amenable to PCI by the interventional cardiologist performing the PCI. Non–IRA lesions were identified as not being responsible for the acute MI when confronted with the infarct territory determined by the diagnostic electrocardiogram (ECG). The full list of the inclusion and exclusion criteria is provided in the Supplementary Appendix.

In both groups, complete revascularization during index procedure was encouraged. If not possible, however, complete revascularization could be performed during another staged procedure, as early as possible, before hospital discharge and no later than 5 days after the initial procedure.

The use of drug-eluting stents was encouraged. Patients of both groups received optimal medical therapy as per the guidelines.^8^

The study protocol was approved by a French national ethics committee. A data monitoring committee provided trial oversight and assessed the safety profile of the trial. Independent clinical research associates monitored the sites and checked the collected data. All events were analyzed and adjudicated by an independent, three-person, blinded clinical evaluation committee. The clinical outcomes at one-year have been reported previously^7^. Three-year follow-up was planned, using the same primary endpoint.

Informed consent was obtained after completion of the procedure on the infarct-related artery; either orally (with subsequent signature) in the case of immediate PCI of the non-infarct-related arteries, or in writing after the initial procedure had been completed in the case of delayed (staged) PCI of the non-infarct-related arteries. Randomization (1:1; in blocks of 2 or 4 randomly permuted) was stratified by center and timing of procedure (immediate or staged) and was performed immediately after obtaining consent, through a web-based centralized system (Cleanweb software, Telemedicine technologies SAS, Boulogne-Billancourt, France).

### FFR measurement

FFR was measured in all lesions judged to have a stenosis of ≥50% by visual estimation using the Radi Medical Systems wire (Abbott). FFR measurement technique is detailed in the protocol. In the FFR group, an FFR value ≤0.80 was considered significant for ischemia with a recommendation that the corresponding PCI be performed. Repeating FFR measurement after completion of PCI was encouraged. An FFR >0.80 was considered non-significant for ischemia and PCI on the corresponding lesion was not to be performed.

### Follow-up

Follow-up was conducted during outpatient clinic visits scheduled at 30 days and at 6, 12, and 36 (±1) months after primary revascularization. Patients for whom no outpatient visit was possible were contacted by mail or telephone. All patients enrolled in the study were monitored for adherence to the protocol and all critical data reported in the case report form were monitored.

### Study outcomes

The primary outcome was the composite of all-cause death, non-fatal MI, and unplanned hospitalization leading to urgent revascularization (major adverse cardiac events, MACE) at one year. We keep this composite endpoint at 3 years as primary outcome for this follow-up study. Secondary outcomes included the following: individual components of the primary outcome; any revascularization (urgent or elective); urgent revascularization for any non-infarct-related artery target lesion; rehospitalization for angina or for acute heart failure; any rehospitalization in a cardiology department; health-related quality of life (as measured by the score on the European Quality of Life–5 Dimensions [EQ-5D] scale);^9^ number of anti-anginal medications. Definitions of the trial outcomes are presented in the Supplementary Appendix.

### Statistical analysis

All analyses were performed on an intention-to-treat basis and all participants were analyzed according to their randomly allocated treatment. Clinical event rates and other categorical data are summarized as percentages. Continuous data are presented as means (standard deviations) or as medians [interquartile range (IQR)].

Kaplan–Meier plots were constructed for time-to-event outcomes, with treatment effects estimated using Cox models and results presented as hazard ratios with 95% confidence intervals (CIs).^10^ A Schoenfeld test was used to check the proportional-hazards assumption. To take into account the fact that trial participants dying early in the trial cannot experience a subsequent nonfatal outcome, we performed sensitivity analyses using Fine and Gray models.^11^ For the numbers of anti-anginal medications at 36 months, a negative binomial model was performed to estimate the mean number of medications in each group. Treatment effect was estimated using the ratio of the two means. All models were adjusted for the timing of the procedure (immediate or delayed; stratification factor).

Planned sub-group analyses of the primary outcome were performed according to age (<65 years vs. ≥65 years), sex, presence of cardiovascular risk factors (diabetes, hypertension, dyslipidemia, family history of coronary artery disease), history of cardiovascular disease, and clinical presentation (Killip class I vs. ≥2). We alsoe performed a sensitivity analysis examining only patients who got an FFR, comparing those in whom FFR was positive in (FFR with ≥ 1 PCI group, n=293) and those with a negative FFR (FFR without PCI group, deferred group, n=193) for the primary outcome at 3-years. Finally, we performed a pooled analysis of published summary data from the FRAME-MI and FLOWER MI trials using the same composite outcomes (death, re-MI or any repeat revascularization) in STEMI patients. The common risk ratio was obtained using a fixed effect Mantel Haenszel model.

A two-sided P value of ≤0.05 for the primary outcome was considered significant. Secondary outcomes are presented with effect-size estimates and 95% CIs. The widths of the CIs have not been adjusted for multiplicity and any inferences drawn from these intervals may not be reproducible. Analyses were performed using SAS software, version 9.2 (SAS Institute, North Carolina, US), R statistical software version 4.0.2 (R Core Team, Vienna, Austria) and RevMan version 5.

## RESULTS

### Participants and follow-up

Between December 18, 2016 and December 6, 2018, 1171 patients with STEMI and MVD were enrolled at 41 sites in France. Of these, 590 were randomly assigned to the FFR-guided group and 581 to the angiography-guided group. The characteristics of the patients at baseline were similar between groups (Table 1).

**Table 1.** Baseline Characteristics of the Patients.*

|  | Total | FFR-Guided PCI | Angio-Guided PCI | P |
| --- | --- | --- | --- | --- |
| Characteristic | (n=1163) | (n=586) | (n=577) | Value |
| Age – yr | 62.2±11.2 | 62.5±11.0 | 61.9±11.4 | 0.37 |
| Median (IQR) | 61.0 (54.0-70.0) | 61.0 (54.3-70.0) | 62.0 (54.0-70.0) |  |
| BMI (Kg/m<sup>2</sup>), † Median (IQR) | 27.1 (24.2-29.4) | 26.7 (24.2-29.1) | 26.6 (24.4-29.7) | 0.36 |
| Missing data | 13/1163 | 6/586 | 7/577 |  |
| Male sex – no. (%) | 966 (83.1) | 498 (85.0) | 468 (81.1) | 0.08 |
| **Medical history** |  |  |  |  |
| Hypertension – no. (%) | 515 (44.3) | 253 (43.2) | 262 (45.4) | 0.44 |
| Diabetes mellitus – no. (%) | 189 (16.3) | 107 (18.3) | 82 (14.2) | 0.06 |
| Hypercholesterolemia – no. (%)‡ | 469 (40.3) | 232 (39.6) | 237 (41.1) | 0.61 |
| Current smoker – no. (%) | 445 (38.3) | 235 (40.1) | 210 (36.4) | 0.33 |
| Family history of CAD – no. (%) | 326 (28.3) | 173 (29.9) | 153 (26.7) | 0.23 |
| Previous myocardial infarction – no. (%) | 76 (6.5) | 45 (7.7) | 31 (5.4) | 0.11 |
| Previous PCI – no. (%) | 103 (8.9) | 59 (10.1) | 44 (7.6) | 0.14 |
| Previous stroke – no. (%) | 33 (2.8) | 16 (2.7) | 17 (3.0) | 0.83 |
| Peripheral arterial disease – no. (%) | 39 (3.4) | 16 (2.7) | 23 (4.0) | 0.23 |
| Chronic renal insufficiency – no. (%)§ | 23 (2.0) | 11 (1.9) | 12 (2.1) | 0.80 |
| Cancer – no. (%)¶ | 40 (3.4) | 23 (3.9) | 17 (3.0) | 0.49 |
| **Location of infarct – no. (%)\*\*** |  |  |  | 0.10 |
| Anterior | 370 (32.2) | 173 (29.8) | 197 (34.6) |  |
| Inferior | 678 (59.0) | 359 (61.9) | 319 (56.0) |  |
| Posterior | 26 (2.3) | 9 (1.6) | 17 (3.0) |  |
| Posterolateral | 67 (5.8) | 36 (6.2) | 31 (5.4) |  |
| Left bundle branch block | 9 (0.8) | 3 (0.5) | 6 (1.1) |  |
| Impossible to determine | 13 (1.1) | 6 (1.0) | 7 (1.2) |  |
| **Arteries with stenosis – no. (%) per patient** |  |  |  | 0.05 |
| 1 | 18/1163 (1.5) | 7/586 (1.2) | 11/577 (1.9) |  |
| 2 | 871/1163 (74.9) | 424/586 (72.4) | 447/577 (77.5) |  |
| 3 | 266/1163 (22.9) | 151/586 (25.8) | 115/577 (19.9) |  |
| Missing data | 8/1163 (0.7) | 4/586 (0.7) | 4/577 (0.7) |  |
| **Killip class ≥ 2 – no./total no. (%)** | 66/1105 (6.0) | 37/556 (6.7) | 29/549 (5.3) | 0.60 |
| LVEF — %, Median (IQR) | 50 (45-60) | 50 (45-58.3) | 50 (45-60) | 0.32 |
| Missing data | 46 | 26 | 20 |  |

\* Plus-minus values are means ± SD. CAD denotes coronary artery disease; FFR, fractional flow reserve; IQR, interquartile range; LVEF, left ventricular ejection fraction; and PCI percutaneous coronary intervention.
† Measurements for BMI (body-mass index; the weight in kilograms divided by the square of the height in meters).
‡ Patients described as having hypercholesterolemia were either receiving treatment with cholesterol-lowering medications or were known to have elevated levels of cholesterol (>200 mg per deciliter [5.2 mmol per liter]).
§ Patients reported as having a clinical history of chronic kidney disease.
¶ Does not include non-melanoma skin cancer.
\*\*The location of the infarct was determined on the diagnostic electrocardiogram.

All sites participated in the 3-year follow-up, with a median duration of 40.7 months (IQR, 36.0 to 46.6) in the FFR-guided group, and 40.3 months (IQR, 36.4 to 46.6) in the angiography-guided group. Information on vital status at 3 years was available in 100% of the patients. Complete follow-up information for other events was available through 3 years for 498 of 586 patients in the FFR-guided group and 502 of 577 patients in the angiography-guided group (Supplementary Appendix Figure S1).

### Procedure results

For non-culprit lesions, staged intervention was used in >95% of the patients in both groups. Mean time delay between interventions was 2.6±1.4 days in the FFR-guided group and 2.7±3.3 days in the angiography-guided group and median time delay was 2 days [2-3] with both strategies. In the FFR-guided group, FFR measurement was attempted in 561 of 586 patients (95.7%), and failed in 13 patients with no severe adverse event (Supplementary Appendix Table S4). PCI was performed in 388 of 586 patients (66.2%). In the angiography-guided group, PCI was performed in 560 of 577 patients (97.1%). The mean number of stents used per patient for non-culprit lesions was 1.01±0.99 with FFR-guided PCI and 1.50±0.86 with angiography-guided PCI.

Medications administered during the procedure, at discharge and at three years are described in Supplementary Appendix Table S5.

### Primary outcome

Clinical outcomes are summarized in Table 2. At three years, the composite primary outcome occurred in 52 patients in the FFR-guided group and in 44 in the angiography-guided group (hazard ratio, 1.19; 95% CI, 0.79 to 1.77; P=0.4) (Figure 1). The lack of benefit of FFR on the primary outcome was consistent across the prespecified subgroups (Supplementary Appendix Fig. S2).

**Figure 1.**
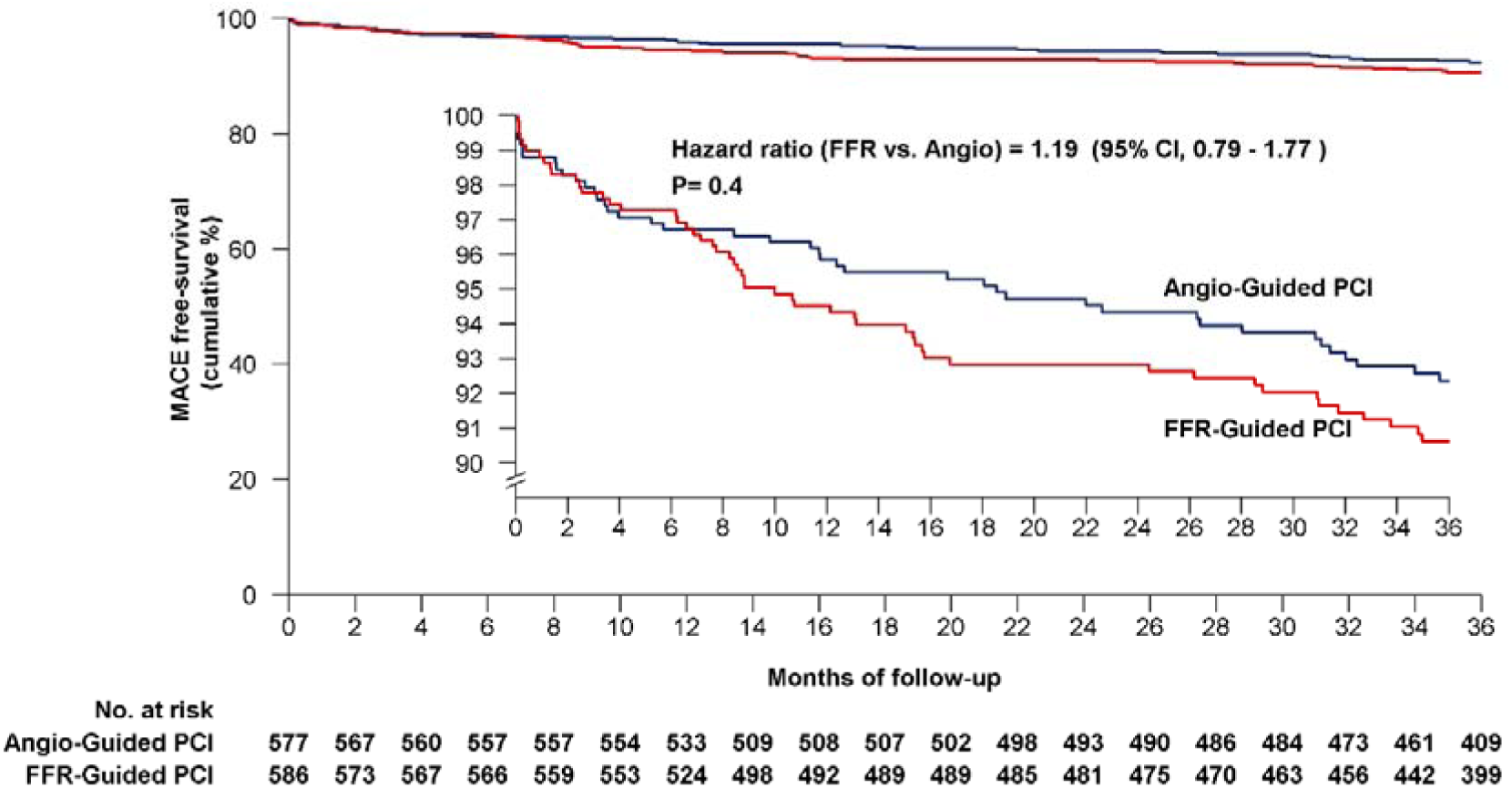
Kaplan–Meier Curves of the Primary Outcome at 3 Years. The primary outcome was a composite of death from any cause, nonfatal myocardial infarction, or unplanned hospitalization leading to urgent revascularization. The inset shows the same data on an expanded y axis.

**Table 2.**
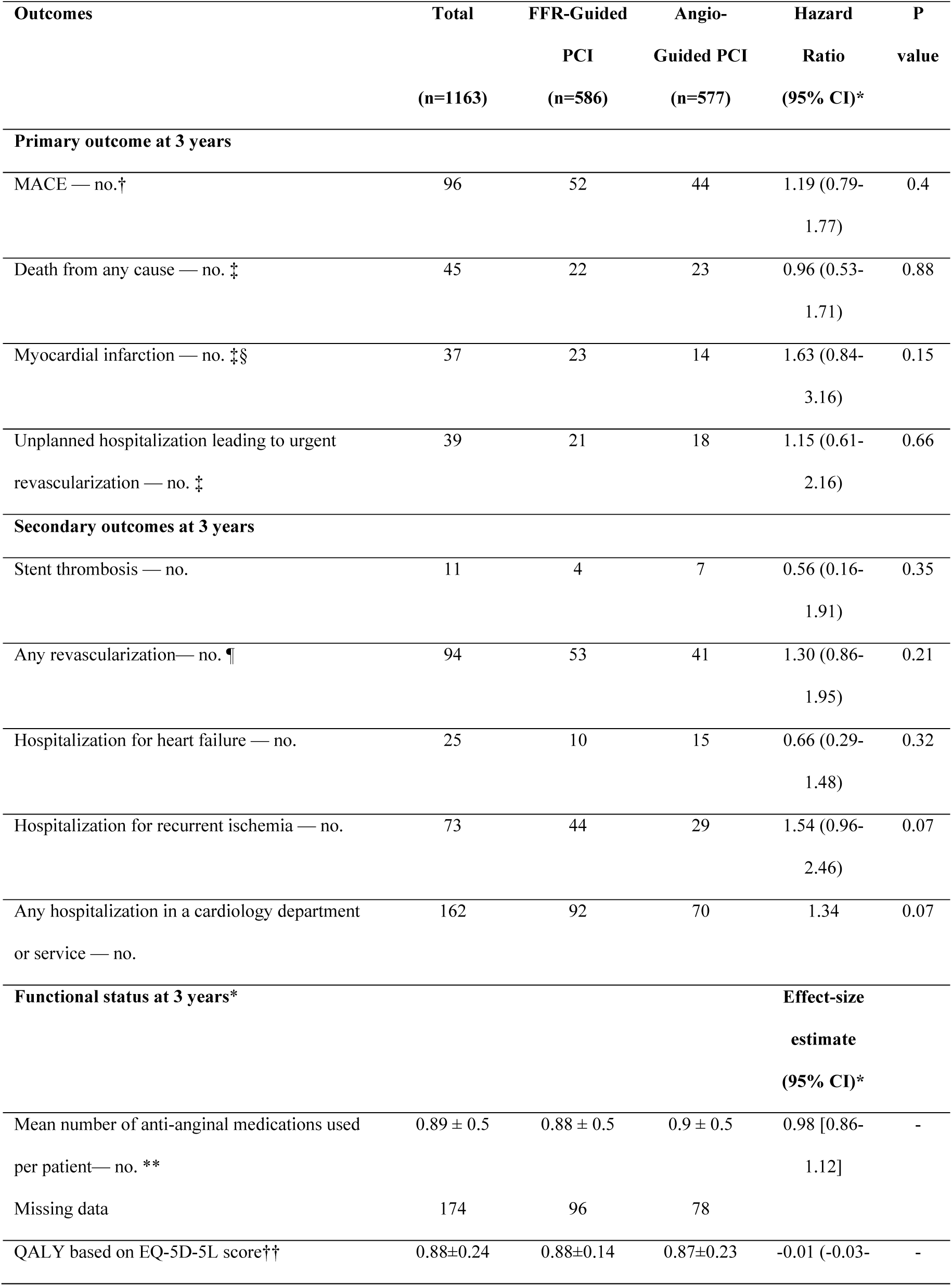
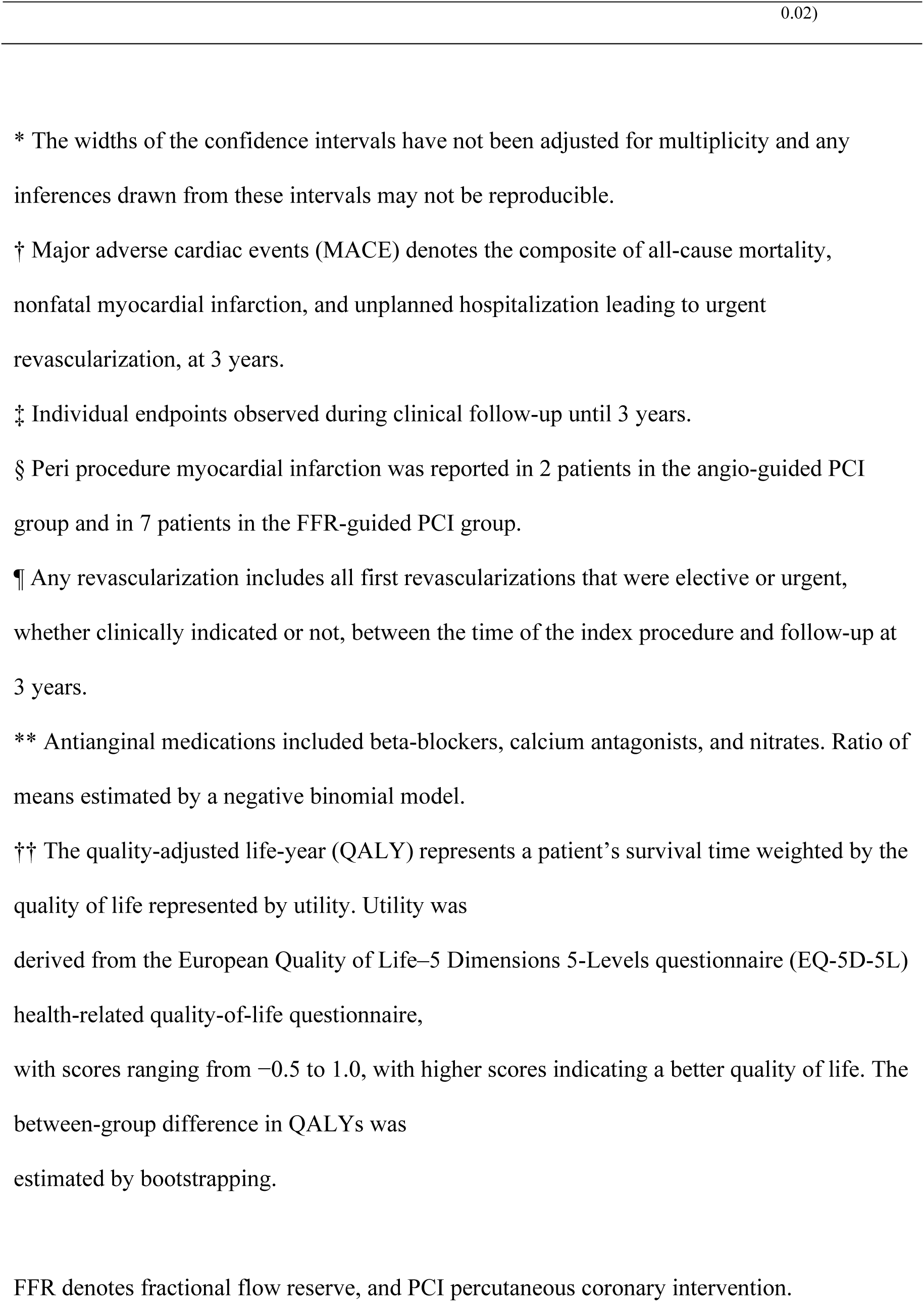
Prespecified Clinical Outcomes at 3 Years.

### Secondary clinical outcomes

Death from any cause occurred in 22 and 23 patients for the FFR-guided and angiography-guided groups, respectively (HR 0.96; 95% CI, 0.53 to 1.71); nonfatal re-infarction in 23 and 14 patients (HR 1.63; 95% CI, 0.84 to 3.16), and unplanned hospitalization leading to urgent revascularization in 21 and 18 patients (HR 1.15; 95% CI, 0.61 to 2.16), respectively. Kaplan-Meier curves for these three components of the primary outcome are shown in Figure 2. Cardiac deaths were reported in 7 patients in the FFR-guided group and in 4 patients in the angiography-guided group (Supplementary Appendix Table S6). For clinical outcomes that did not include death, competing-risks analyses were performed, which showed similar results to the main analyses (Supplementary Appendix Table S7).

**Figure 2.**
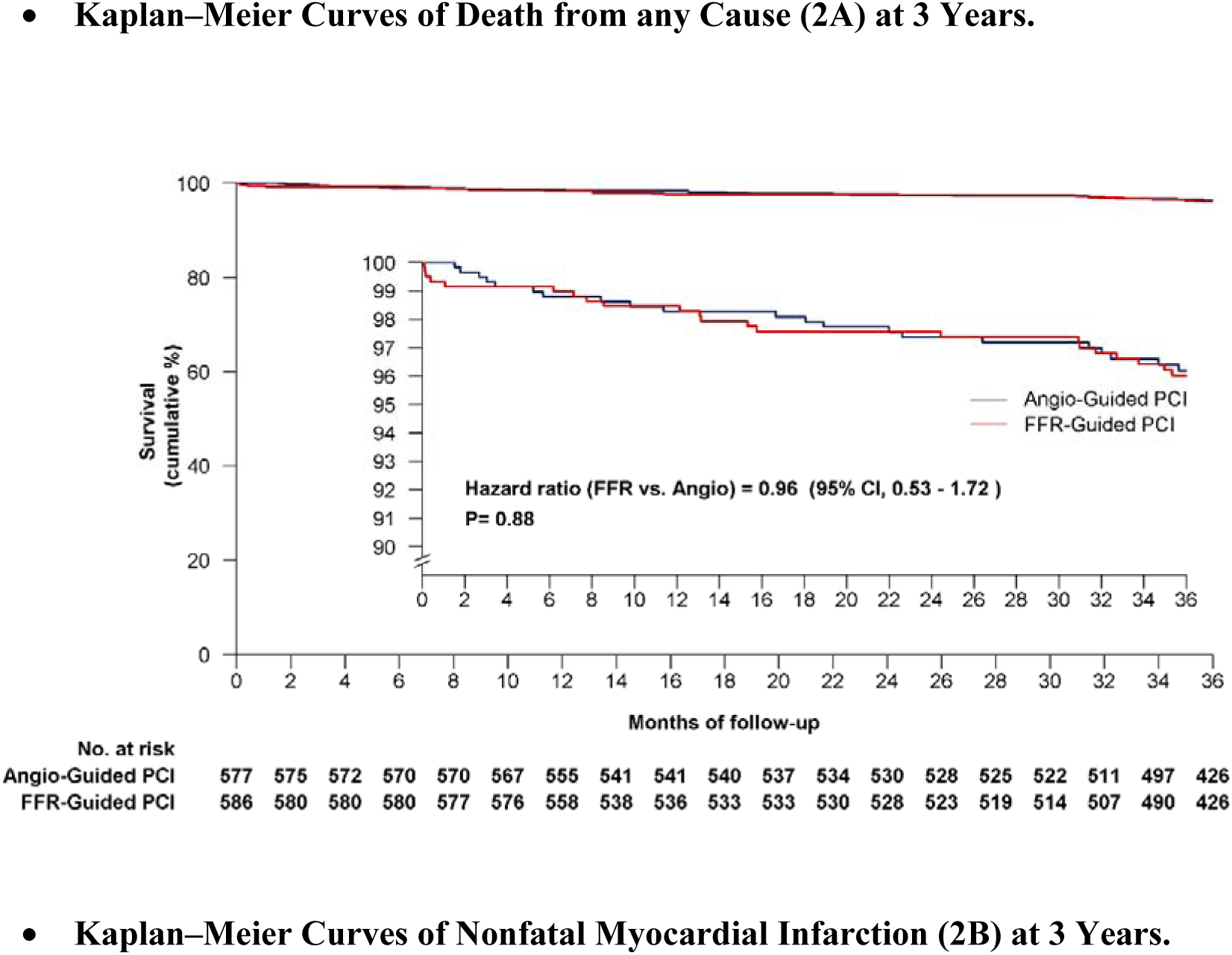
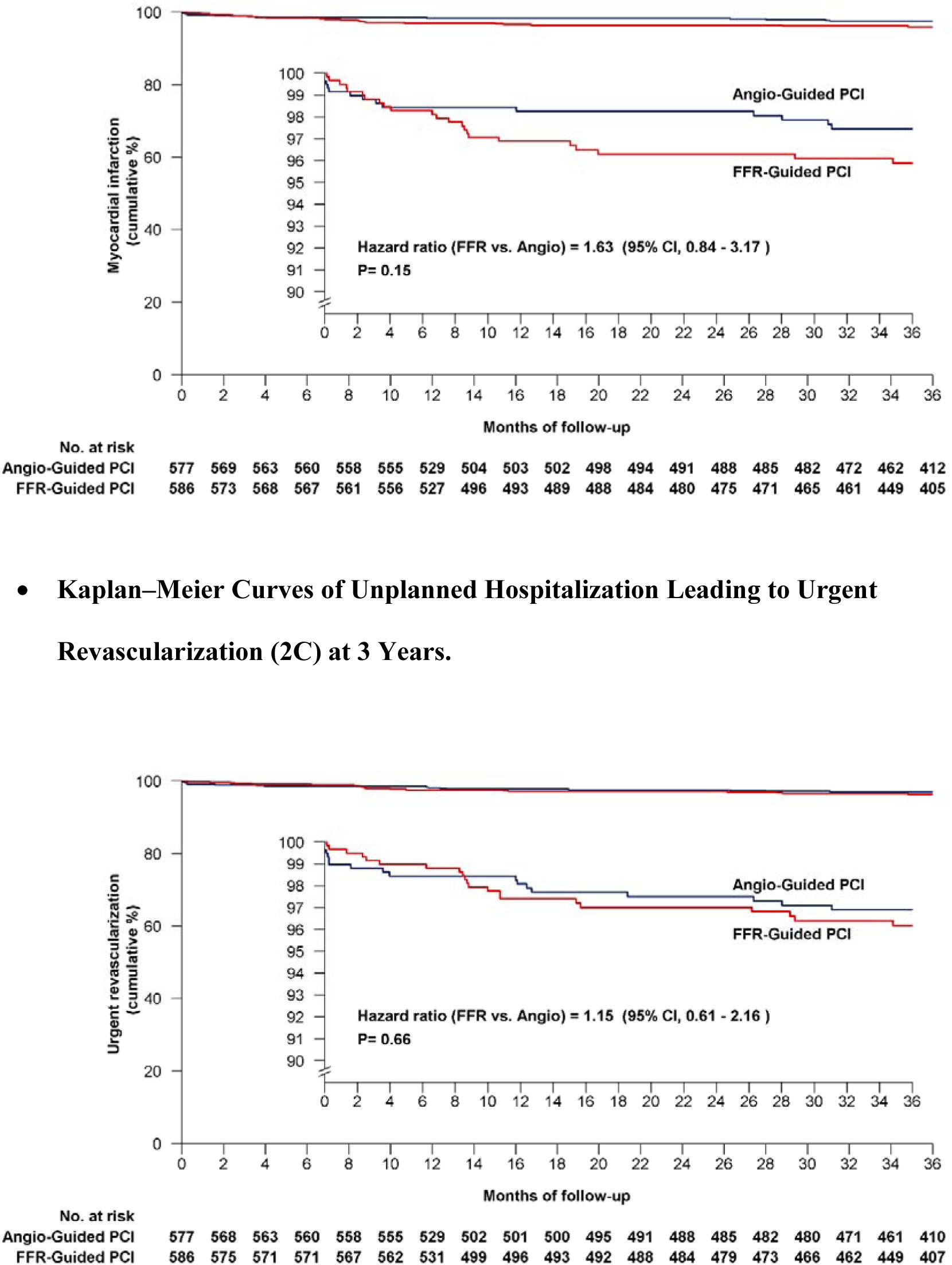
**Kaplan–Meier Curves of Death from any Cause (2A), Nonfatal Myocardial Infarction (2B), and Unplanned Hospitalization Leading to Urgent Revascularization (2C) at 3 Years.**

At three years, the composite of death from any cause, nonfatal re-infarction or any revascularization occurred in 76 patients in the FFR-guided group and in 65 in the angiography-guided group (HR, 1.18; 95% CI, 0.85 to 1.64) (Supplementary Appendix Figure S3).

### Clinical outcomes in patients treated medically versus patients receiving at least one PCI for non-culprit lesions within the FFR arm

In the FFR-guided group, the primary outcome occurred in 31 of the 388 patients who had undergone ≥ 1 PCI versus 21 of the 198 patients without PCI (HR 0.72; 95% CI, 0.42 to 1.26, for performing vs deferring PCI; Supplementary Appendix Table S8).

## DISCUSSION

Among patients with STEMI and MVD, an FFR-guided strategy was not superior to an angiography-guided strategy for reducing the risk of the composite of death from any cause, non-fatal MI, and unplanned hospitalization leading to urgent revascularization at three years. The individual components of the primary outcome, as well as all other clinical outcomes, did not differ significantly between the two groups. These results are consistent with those observed at 1 year of follow-up.

The results of our trial are not in agreement with those of the FRAME-AMI trial comparing FFR-guided vs angiography guided PCI in acute MI (either STEMI for 265 patients or NSTEMI for 297 patients) and MVD, which showed that an FFR-guided PCI reduced a composite of death, MI or repeat revascularization at a median follow-up of 3.5 years. The reduction in the composite end-point was driven mainly by a significantly lower risk of death and MI in the FFR-guided PCI group. Direct comparison of the two trials results is difficult due to different patient populations. FLOWER MI enrolled only patients with STEMI whereas the FRAME-AMI trial enrolled patients with either STEMI or NSTEMI. Consequently, the FRAME-AMI study population was at higher cardiovascular risk with a higher rate of patients with 3-vessel disease. Indeed, in the FRAME-AMI trial, the results appeared to be driven mostly by the NSTEMI population (27 primary outcome events vs. 9 events) whereas the difference was much smaller in the STEMI population (13 vs. 9 events) suggesting that for patients with STEMI the results of FLOWER-MI and FRAME-AMI may not be all that different. In the FLOWER-MI trial where most patients underwent staged non-culprit lesion PCI, there was no appreciable increase in peri procedural events in the angiography-guided PCI group (0.3% vs 1.9%). In contrast, in the FRAME-AMI trial, where 60% underwent an immediate multivessel PCI, there was a sharp early increase in events in the angiography-guided PCI group (4% vs 1.1%). The reduction in all-cause mortality observed in the FRAME-AMI trial between FFR-versus angiography-guided strategies for complete myocardial revascularization is huge; such a large difference has not been reported in previous trials comparing complete versus incomplete revascularization at the acute stage of STEMI such as the Complete versus Culprit-Only Revascularization Strategies to Treat Multivessel Disease after Early PCI for STEMI (COMPLETE) trial and should therefore be interpreted with caution.^12,13^ Finally, a pooled analysis of the results from the two trials using the same composite outcomes (death, re-MI or any repeat revascularization) in STEMI patients shows no statistically significant benefit of the FFR-guided strategy over the angiography-guided strategy (risk ratio 1.11; 95% CI 0.83-1.48) (Figure 3).

**Figure 3.**
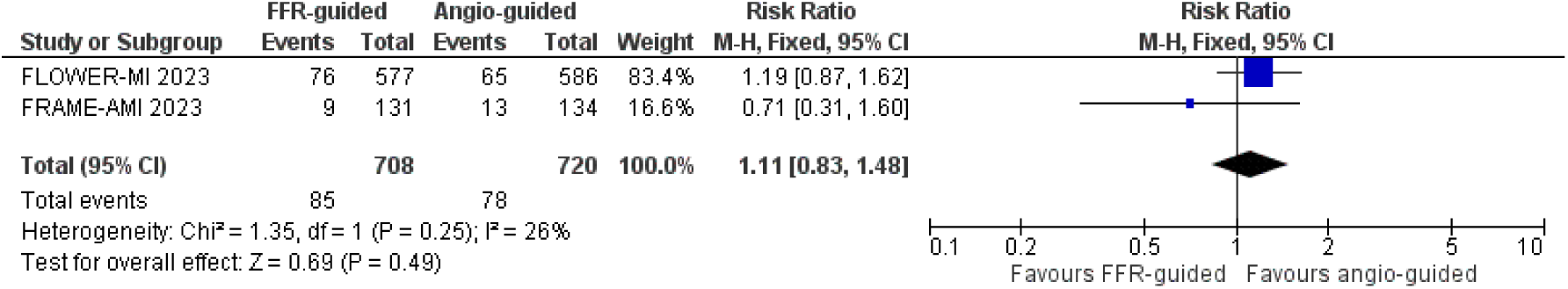
Pooled analysis of FFR-guided versus angio-guided PCI in STEMI patients with multivessel disease. Composite outcomes (death, re-MI or any repeat revascularization) in the FRAME-AMI and FLOWER-MI trials.

The physiology-guided approach for the functional assessment of lesion severity is well established in patients with chronic coronary syndromes (CCS) where it has been shown to be superior to an angiography-guided strategy.^14,15^ In the setting of acute coronary syndromes (ACS), the role of a physiology-guided strategy is much less clear because, while in CCS most plaques are fibro-calcific with a low propensity to plaque rupture, in ACS about one-half of all plaques are inflamed lipid-rich thin-capped fibroatheromas with a higher propensity for plaque rupture.^16–18^ The fate of these non-culprit lesions and their clinical consequences may be more closely linked to plaque morphology than to their physiological significance, implying that deferral of some physiologically insignificant lesions with high-risk morphological features such as inflammation and thin-capped fibroatheromas could still be associated with increased cardiovascular events. Recently, a meta-analysis by Liou et al has shown that the event rate in ACS patients was much higher than in patients with CCS despite following an FFR-guided revascularization strategy.^19^

The limitations of the present analysis are the same as those reported in the 1-year follow-up analysis of the FLOWER MI trial. First, with only 96 primary outcomes events at three years, the trial was underpowered compared with the assumptions we made when the trial was designed, and we calculated the sample size. Given the wide CIs for the estimate of effect, the findings do not allow for a definitive interpretation. In the setting of STEMI patients, assessing peri-procedural MI (which represents one of the benefits of FFR in chronic coronary syndromes) is difficult and we cannot exclude that this event may have been under-reported.

In conclusion, in patients with STEMI and MVD undergoing PCI, we found no significant benefit of an FFR-guided strategy as compared with an angiography-guided strategy in the management of non-culprit lesions with respect to the risk of death, MI, or urgent revascularization at one and 3 years following the index event.

## SUPPLEMENTARY DATA

Supplementary data are available at *Circulation* online.

## Trial Registration

FLOWER-MI ClinicalTrials.gov number, NCT02943954

## NONSTANDARD ABBREVIATIONS AND ACRONYMS

ACS: acute coronary syndrome
CCS: chronic coronary syndrome
COMPLETE: Complete versus Culprit-Only Revascularization Strategies to Treat Multivessel Disease after Early PCI for STEMI
FFR: fractional flow reserve
FLOWER-MI: Flow Evaluation to Guide Revascularization in Multivessel ST-Elevation Myocardial Infarction
MVD: multivessel disease
FRAME-AMI: Fractional Flow Reserve vs. Angiography-Guided Strategy for Management of Non-Infarction Related Artery Stenosis in Patients with Acute Myocardial Infarction
IRA: infarct-related coronary arteries
MACE: major adverse cardiac events
PCI: percutaneous coronary intervention
STEMI: ST-elevation myocardial infarction

## ACKNOWLEDGEMENTS

The authors are deeply indebted to all patients who accepted to participate in the surveys, and to the physicians who took care of the patients at the participating institutions. The authors would like to thank Shohreh Azimi, Laura Le Mao, Aurelia Dinut, Hakima Manseur, Hasina Rakotosamimanana, Axel Bouffier, Hajar Chouiki, Lynda Arab, Nivetha Atputhanathan, Hasina Rabetrano, Odélia Bitton, Sabrina Boudif, Salima Bouslah, Myriam Calvet, Alexandre Cuquemelle, Meriem Damouche, Sarra Elouaret, Sabrina Fiori, Muriel Gernet, Fatiha Khedim, Rana Korbi for their help in managing the FLOWER-MI study.

## DATA AVAILABILITY

The FLOWER MI trial is planning to continue analysis, including post hoc subgroup analysis. Any relevant inquiry should be emailed to Dr. Etienne PUYMIRAT

